# Genomic profiling implicates candidate genes and mutagenic pathways driving lung cancer recurrence

**DOI:** 10.64898/2025.12.24.25342926

**Authors:** Laura Luhari, Ann Valter, Alexander T Bahcheli, Kevin CL Cheng, Masroor Bayati, Anu Ustav, Agne Velthut-Meikas, Kersti Oselin, Jüri Reimand

## Abstract

Lung cancer remains the leading cause of cancer-related deaths worldwide, with tumor recurrence a major contributor to its high mortality. The genetic and molecular mechanisms of recurrence remain poorly understood. Using whole-exome sequencing of 155 primary non-small cell lung cancers, we studied the mutational landscape and driver alterations associated with recurrence. Primary tumors that developed recurrence had higher mutational burden, including hypermutated tumors explained by mutations in DNA polymerase or mismatch repair pathways. Mutational signatures of reactive oxygen species were associated with recurrence. Combined mutations in *TP53* and *CDKN2A* were enriched in non-recurrent tumors, while *ATRNL1* mutations were enriched in recurrent tumors. Pathway analyses implicated DNA repair and cilium organisation processes with tumor recurrence and highlighted 50 additional candidate genes including *BRCA2*. Recurrence-associated genes showed essentiality in lung cancer cell lines and included known therapeutic markers, indicating their functional and translational relevance. This analysis provides insights into the molecular basis of lung cancer recurrence and informs experiments to develop diagnostic and therapeutic strategies.

## INTRODUCTION

Lung cancer is the leading cause of cancer-related mortality that accounted for 2.4 million new cases and 1.8 million deaths worldwide in 2022 [1]. Lung cancer etiology is multifactorial and includes tobacco exposure, environmental pollutants, occupational carcinogens, genetic susceptibility, and chronic pulmonary diseases. Non-small cell lung cancer (NSCLC) is the predominant form of lung cancer that includes the adenocarcinoma and squamous cell carcinoma subtypes with markedly distinct biological profiles and etiology [2].

Recurrence of lung cancer is a major clinical challenge as many patients experience disease relapse within two years after treatment [3,4]. Recent developments in targeted therapies and immunotherapies have substantially improved survival rates in early-stage NSCLC [5] and driver genes such as *KRAS*, *EGFR*, *ALK*, *BRAF*, and *RET* encode established therapeutic targets [6], underlining the value of tailoring treatment strategies to patients based on the genetic characteristics of their disease. Despite these advancements, the five-year overall survival of NSCLC is only around 33-77% and limited therapeutic options are available for recurrent disease. Further, recurrence remains the primary cause of death from lung cancer [7], highlighting the urgent need for improved strategies to predict, monitor, and prevent disease relapse.

Histopathological features and the extent of disease spread at the time of diagnosis are established prognostic factors for NSCLC; however, additional biomarkers are emerging [8]. Genomic and multi-omic profiling of cancer samples is a key approach to deciphering the genomic drivers, molecular biomarkers and potential therapeutic targets of NSCLC [9,10]. Lung cancer evolution is driven by somatic mutations in oncogenes and tumor suppressor genes such as *EGFR, PTEN, TP53, RB1, CDKN2A,* and *PIC3CA.* Tumors can acquire resistance-enabling mutations in response to targeted therapies, such as *EGFR* mutations that have been found to emerge during targeted therapy [11]. Further, lung cancers have high intratumoral heterogeneity and comprise subpopulations of cells characterised by distinct sets of genomic alterations, whereas certain alterations acquired in early evolution ultimately read to recurrence and metastasis [12]. Thus, genomic features already present in primary, untreated tumors may promote later recurrence.

The genetic mechanisms of lung cancer recurrence remain poorly understood. While prior studies have investigated genomic driver alterations or transcriptomic signatures associated with lung cancer recurrence, well-defined targetable pathways or biomarkers have not yet emerged. Multivariable analysis of molecular signatures in NSCLC revealed several genomic drivers as among the independent prognostic factors of disease-free survival [13]. A systematic analysis of early-stage NSCLCs in the Cancer Genome Atlas (TCGA) cohort reported a recurrence-associated gene signature comprising *ATR, ERBB2, KDR and MUC6* gene mutations, *ROS1* and *NTRK1* fusions and alterations in VEGF signalling pathway [14]. Another analysis of NSCLC samples in TCGA derived a gene signature to estimate recurrence risk [15]. To our knowledge, prior studies have not investigated genetic alterations in lung cancer recurrence by directly comparing early-stage primary lung cancers that subsequently developed recurrent or non-recurrent phenotypes.

Here we aimed to identify genomic alterations in genes and molecular pathways associated with lung cancer recurrence. Using a comprehensive analysis of exome sequencing data from a cohort of patients with primary NCSLCs that later developed recurrence or remained disease-free, we revealed aspects of the mutational landscape, candidate driver genes and molecular pathways that were associated with tumor recurrence. The candidate genes and molecular pathways outlined in this study may contribute to a better understanding of the foundational biology of lung cancer recurrence and provide insights for therapy and biomarker development.

## MATERIALS AND METHODS

### Cohort

155 patients were selected from North Estonia Medical Centre (NEMC) Thoracic Oncology Database based on development of lung cancer recurrence (n = 72) or no recurrence (n = 83) as matched for treatment, histology, age, sex, and follow-up time. The study was approved by the ethics committee of Estonian National Institute for Health Development (#KK2453; 2018-09-29). Informed consent was waived due to minimal or no patient risk involved.

### Whole exome sequencing (WES)

DNA extraction from FFPE tumor samples, quantification and library generation were performed using standard protocols (**Supplementary Methods**). Tumor samples were sequenced using 150-bp paired-end sequencing (average 734x) on Illumina NovaSeq 6000 at Intermountain Precision Genomics (Utah, US). Germline reference samples were not available. Reads were aligned to GRCh38 and consensus SNVs and indels were called from three methods (Mutect2, Strelka2, FreeBayes). Germline variants were filtered using a panel of normals from the gnomAD database (v4.0).

### Statistical associations of genomic features

Standard statistical tests were used to compare mutation burden, clinical stages, histology, and protein-coding mutations in polymerase (*POLE*, *POLD1*, *POLQ, POLE2*) or mismatch repair genes (*MSH2*, *MSH6*, *MLH1*, *PMS2*) between R and NR tumors. Tumors with >3000 SNVs or indels were considered hypermutated. Non-parametric Wilcoxon rank-sum tests were used for numeric features and hypergeometric tests were used for binary features. Statistically significant associations were highlighted (P < 0.05).

### Mutational signatures

Single base substitution (SBS) signatures were mapped to samples using SigProfilerAssignment [16] and COSMIC V3 catalogue [17], excluding artefact signatures. SBS contributions between R and NR tumors were compared using Two-sided Wilcoxon rank-sum tests and adjusted using false discovery rate (FDR).

### Gene-based analyses

Mutations in each gene were evaluated between R and NR tumors excluding hypermutated tumors and using hypergeometric tests, with resulting P-values adjusted using FDR. First, we tested known NSCLC driver genes [18]. Second, we tested all genes with ≥10 protein-coding mutations in two tiers (FDR < 0.05; P < 0.01). Combined mutations in *TP53* and *CDKN2A* were tested separately.

### Pathway-based and functional analyses of candidate genes

Pathway enrichment analysis was performed on gene P-values from above separately for R- and NR-enriched genes using ActivePathways [19] (adjusted P < 0.1) (**Supplementary Methods**). Next, we studied rank-normalised gene expression of top five candidate genes in lung cancer samples from TCGA [20] and normal lungs from GTEx [21] using custom permutation tests. Lastly, we examined gene essentiality in NSCLC cell lines in the DepMap database [22], comparing essentiality scores for each gene to reference points (0, potentially essential; -0.5, strongly essential) using one-tailed one-sample Wilcoxon signed-rank tests adjusted with FDR. Lastly, therapeutic associations from OncoKB [23] were examined.

Additional details on Materials and Methods are available in **Supplementary Information**.

## RESULTS

### Exome-wide sequencing of primary lung cancers to study tumor recurrence

To study genomic alterations associated with lung cancer recurrence, we assembled a retrospective cohort of 155 primary lung tumors (**Table 1**). The cohort included 72 patients in the recurrence (R) group that developed recurrence during a median follow-up period of 1.5 years (95% range: 0.13—7.6 years), and 83 patients in the non-recurrence (NR) group that remained recurrence-free during a median follow-up of 8.5 years (95% range: 4.4—17 years). Major histological subtypes and tumor stages were matched between R and NR groups (**Figure 1A**). Patients received standard curative treatments prior to sequencing that did not include targeted therapies or immunotherapies. Whole-exome sequencing (WES) of tumor samples followed by consensus variant calling from multiple variant-calling tools and panel-of-normals germline variant filtering identified 184,913 single-nucleotide variants (SNVs) and 3043 insertions-deletions (indels), corresponding to an average of 1193 SNVs and 20 indels per tumor and an 88% reduction from the pre-filtered variant set. We note that mutation burden in our cohort appears higher due to incomplete filtering of rare germline variation.

**Figure 1.**
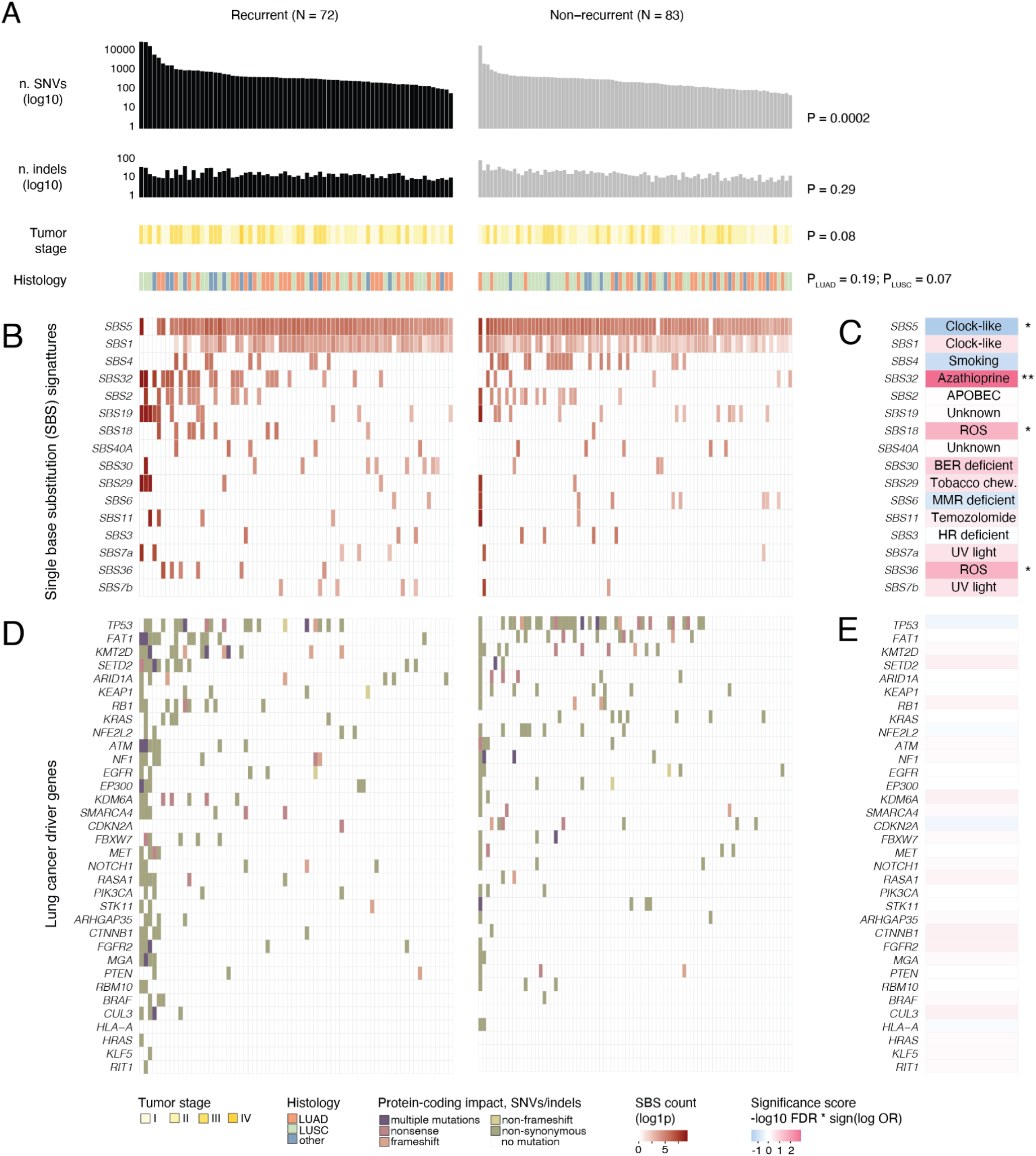
Mutational landscape of primary lung cancers with subsequent recurrent (R) and non-recurrent (NR) outcomes. We profiled 155 lung cancer samples (R = 72, NR = 83) using whole exome sequencing. **A.** Barplot shows tumor mutation burden measured as the total number of single-nucleotide variants (SNVs) and small insertions-deletions (indels). Clinical annotations shown below include tumor stage (I-IV) and histology (adenocarcinoma, squamous cell carcinoma, other). P-values comparing R and NR tumors are shown on the right (Wilcoxon tests for mutation burden and tumor stage, hypergeometric tests for histology). **B.** Quantification of mutational signatures of single base substitutions (SBS) across R and NR tumors. **C.** P-values comparing SBS signature frequencies in R and NR tumors. Pink tiles reflect enrichments of SBS signatures in R tumors and blue tiles reflect enrichments of signatures in NR tumors. P-values were calculated using two-sided Wilcoxon rank-sum tests and adjusted for multiple testing using the false discovery rate (FDR) method. Asterisks indicate statistical significance (* FDR < 0.05, ** FDR < 0.01, *** FDR < 0.001). **D.** Mutations found in established NSCLC driver genes in R and NR tumors. Colors reflect the type of protein-coding mutation. **E.**. Comparison of mutations in established NSCLC driver genes in R and NR tumors. Mutational enrichments in R tumors (pink) or NR tumors (blue) are reflected by the color, and the significance is reflected by the shade. P-values were calculated with two-sided hypergeometric tests and adjusted for multiple testing. Acronyms: lung squamous cell carcinoma (LUSC); lung adenocarcinoma (LUAD); apolipoprotein B mRNA editing enzyme (APOBEC); reactive oxygen species (ROS); base excision repair (BER); DNA mismatch repair (MMR); homologous recombination (HR); ultraviolet (UV);

**Table 1.**
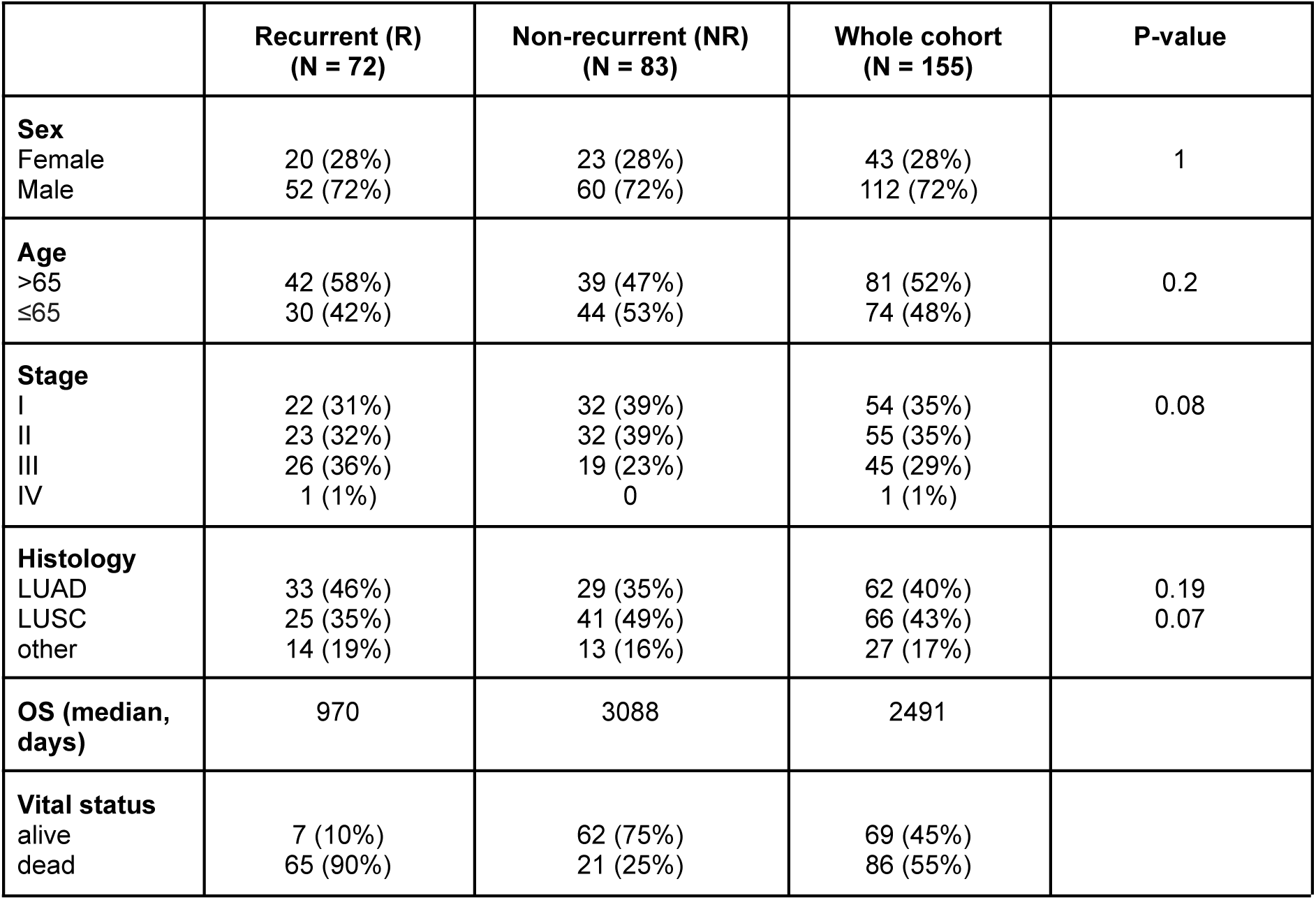
Clinical and pathological characteristics of patients with recurrent (R) and non-recurrent (NR) lung cancer in the study cohort.

### Mutational features of tumors with recurrence phenotypes

First, we examined the overall mutational landscape of our cohort in the context of tumor recurrence. Tumors in the R group displayed a significantly higher SNV mutation burden compared to NR tumors (P = 0.0002, two-tailed Wilcoxon rank-sum test) while indel burden was similar between the groups (P = 0.29) (**Figure 1A**). Six tumors (3.9%) had a hypermutated phenotype that we defined as above 3000 SNVs and indels, in total accounting for 63.8% of mutations across our cohort.

To explore mutational processes in the context of tumor recurrence, we evaluated mutational signatures of single base substitutions (SBS). The most common SBS signatures were associated with aging and clock-like mutagenesis (SBS5), methylcytosine deamination (SBS1), tobacco smoking (SBS4), azathioprine exposure (SBS32), and APOBEC activity (SBS2) (**Figure 1B-C, Table S1**). R tumors were enriched in SBS signatures SBS18 and SBS36, which reflect mutagenesis from reactive oxygen species (ROS) (both FDR = 0.04). The signature SBS32, which is associated with immunosuppression treatment, was also enriched in R tumors (FDR = 0.02), however it was predominantly found in a few hypermutated tumors. NR tumors were also enriched in mutational signatures, including the clock-like SBS5 associated with patient age (FDR = 0.04) and a sub-significant trend with the tobacco smoking signature SBS4 (FDR = 0.1). Collectively, this analysis highlights distinct mutational processes associated with recurrence phenotypes of lung cancer.

Next, we asked if mutations in established cancer driver genes were associated with tumor recurrence. We focused on 31 NSCLC driver genes from the TCGA PanCanAtlas study [18], aiming to find genes in which protein-coding mutations were enriched in R or NR tumors. No significant associations were found at pre-defined significance cutoffs, both when analysing all tumors and after excluding hypermutated samples (hypergeometric tests, FDR < 0.05, **Figure 1D-E**, **Table S2**). Overall, *TP53* was the most frequently mutated gene altered in 32% of tumors, followed by *FAT1*, *KMT2D*, and *SETD2*. In summary, mutational analysis of primary NCSLCs in the context of recurrence indicated differences in mutational processes, while mutations in established driver genes were less informative.

### DNA polymerase and mismatch repair mutations link to tumor recurrence

We investigated tumor recurrence in the context of elevated mutagenesis and associated pathways. First, highly mutated lung tumors often showed mutations in DNA polymerase genes (*POLE*, *POLD1*, *POLE2*, *POLQ*) or mismatch repair (MMR) pathway genes (*MSH2*, *MSH6*, *MLH1*, *PMS2*), consistently with prior evidence [24][25] (**Table S3**). R tumors were enriched in mutations in POL genes combined (20 R *vs*. 5 NR tumors, two-tailed hypergeometric P = 0.0003), MMR genes combined (11 R *vs*. 4 NR tumors, P = 0.03, **Figure 2A-B**), and several genes individually (POLE2, P = 0.003; POLQ, P = 0.006). The six hypermutated tumors each had multiple mutations in POL and MMR genes. Several variants were annotated as pathogenic in the ClinVar database [26]: *POLE* p.E321X truncates the exonuclease domain and likely compromises polymerase proofreading activity, while p.W62X likely causes early loss of function. In *MSH2*, p.Q170X is a pathogenic variant in Lynch syndrome. Additional variants of unknown significance found in these genes are more challenging to interpret functionally.

**Figure 2.**
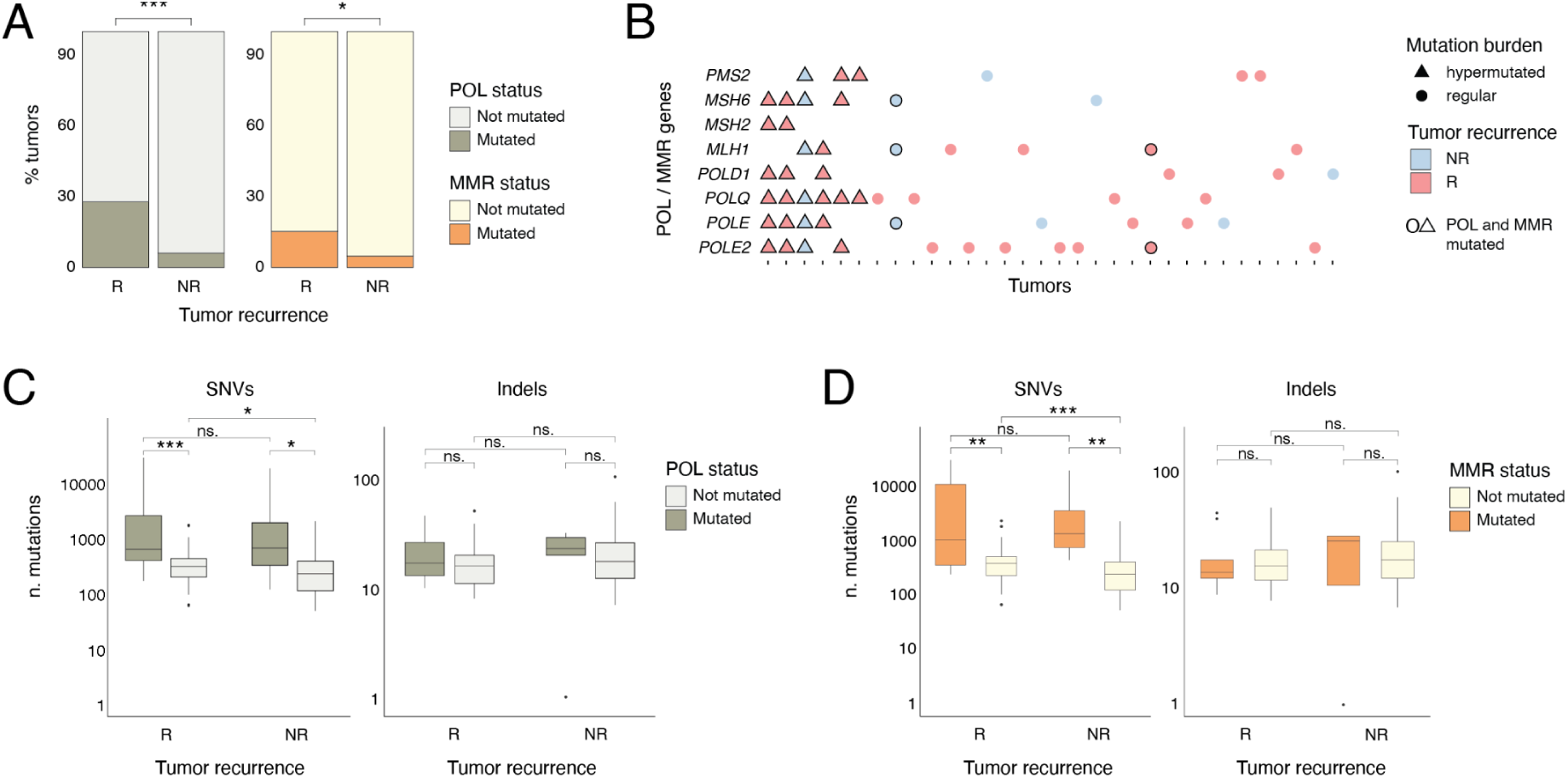
Mutations in DNA polymerase (POL) and mismatch repair (MMR) genes are associated with tumor recurrence and mutation burden. **A.** Associations of tumor recurrence and mutation status of POL or MMR genes. Proportions of R and NR tumors grouped by mutation status in POL or MMR genes, based on protein-coding mutations in POL (*POLE2*, *POLE*, *POLD1*, or *POLQ*) or MMR (*MSH2*, *MSH6*, *PMS2*, *MLH1*) genes, respectively. P-values from two-tailed hypergeometric tests are shown. **B.** Distributions of POL and MMR gene mutations in R and NR tumors. Hypermutated samples (triangles) often have mutations in multiple genes, including a subset of tumors with mutations in both POL and MMR genes (black outlines). **C-D.** Mutation burden in R and NR tumors based on mutation status of either POL genes (panel C) or MMR genes (panel D). SNV burden (left) and indel burden (right) are shown separately. P-values were computed using two-sided Wilcoxon rank-sum tests and asterisks indicate statistical significance (* P < 0.05, ** P < 0.01, *** P < 0.001).

Next, we examined POL and MMR gene alterations in the context of tumor mutation burden. Mutations in POL genes were associated with increased SNV burden both in R and NR tumors (P = 0.0003 and P = 0.047, respectively, two-tailed Wilcoxon rank-sum test, **Figure 2C**), and similar observations were found for MMR genes (**Figure 2D**). This highlights an association between DNA repair defects and hypermutation phenotypes in our cohort. When considering tumors lacking mutations in either POL or MMR genes, R tumors still had higher SNV burden compared to NR tumors (P = 0.01, **Figure 2C-D**), suggesting that additional mutational processes contribute to elevated mutagenesis. No differences in SNV burden were observed between R and NR tumors that carried mutations in POL or MMR genes, indicating that NR tumors may also have high mutation burden when these DNA repair pathways are altered. In contrast to SNVs, tumor indel burden was not associated with mutation status of POL or MMR genes. Our data suggest that primary lung cancers with subsequent recurrence outcomes are more likely to have hypermutation phenotypes caused by mutations in DNA polymerase or mismatch repair genes.

### Candidate genes associated with lung cancer recurrence

We focused again gene mutations in the context of lung cancer recurrence. While no high-confidence associations with tumor recurrence and established driver mutations were found, both *TP53* and *CDKN2A* mutations showed trending enrichments in NR tumors (P = 0.078 and P = 0.074, respectively). Investigating combinations, we found that mutations in either *TP53* or *CDKN2A* were often found in NR tumors (n = 36) compared to R tumors (n = 16), more than expected from chance alone (P = 0.015, two-tailed hypergeometric test) (**Figure 3A**).

**Figure 3.**
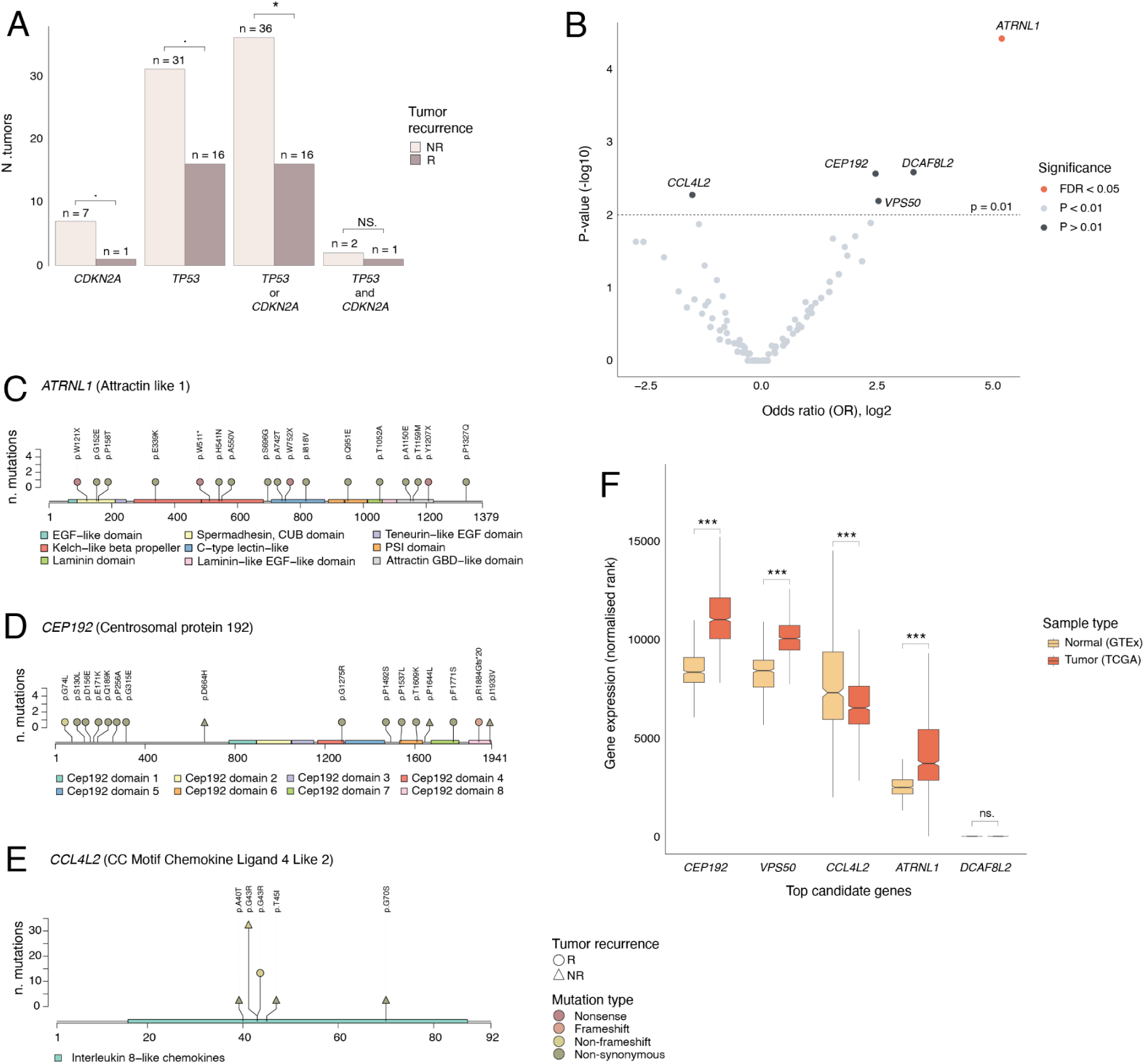
Mutational enrichments in genes associated with lung cancer recurrence. **A.** Mutational frequencies of *TP53* and *CDKN2A* in recurrent (R) or non-recurrent (NR) tumors. Combination of mutations in *TP53* or *CDKN2A* was significantly enriched in NR tumors, while mutations in the two genes alone showed positive trends in NR tumors. P-values were computed using two-tailed hypergeometric tests. **B.** Analysis of mutational enrichments in 252 protein-coding genes shown as a volcano plot. Genes with mutations in at least ten tumors were analysed. FDR-adjusted P-values from two-tailed hypergeometric tests are shown. The top gene *ATRNL1* is printed in red (FDR < 0.05) and four second-tier genes are printed in black (unadjusted P < 0.01). **C-E.** Mutations in candidate genes. Lollipop plots show mutated protein sequence positions (X-axis) and number of mutations (Y-axis). Node shapes reflect outcome (R *vs.* NR) and colors reflect protein-coding mutation impact. **C.** Comparison of gene expression levels of candidate genes in lung tumors from TCGA and normal lung samples from GTEx. Gene expression values were rank-normalised across protein-coding genes and normal and cancer tissues were compared using permutation tests. Asterisks indicate statistical significance (• P < 0.1, * P < 0.05, ** P < 0.01, *** P < 0.001).

Next, we extended our search to 252 protein-coding genes, excluding hypermutated tumors and less-frequently mutated genes (n < 10 patients) to improve statistical power and reduce biases. We found one significant gene at a predefined cutoff (FDR < 0.05): *ATRNL1* was exclusively mutated in tumors with subsequent recurrence (12 R *vs*. 0 NR tumors, P = 3.8 x 10^-5^, FDR = 0.01) and included 13 missense SNVs and four nonsense SNVs (**Figure 3B-C, Table S4-5**). *ATRNL1* (Attractin Like 1) encodes a poorly studied transmembrane protein potentially involved in GPCR signaling. Four additional genes showed sub-significant associations with tumor recurrence (unadjusted P < 0.01; **Figure 3B**). Mutations in *CEP192* were enriched in R tumors (13 R *vs*. 3 NR tumors, P = 0.003, **Figure 3D**). Another gene *CCL4L2* was more frequently mutated in NR tumors (35 NR *vs*. 14 R tumors, P = 0.005). Interestingly, *CCL4L2* mutations form a mutational hotspot for amino acid substitution p.G43R (**Figure 3E**). *CCL4L2* encodes a poorly studied cytokine protein whose function in cancer remains unexplored. Two additional trending associations with tumor recurrence included *VPS50* (10 R *vs*. 2 NR tumors, *P* = 0.006) and *DCAF8L2* (10 R *vs.* 1 NR tumors, P = 0.003) (**Figure S1**). Multiple-testing adjustments indicate that second-tier candidate genes include a few false positives (FDR < 0.33).

Next, we examined expression patterns of the candidate genes, asking if these were differentially expressed between lung tumors and normal lung tissues, using transcriptomics data from TCGA [20] and GTEx [21], respectively. *CEP192*, *VPS50*, and *ATRNL1* were significantly upregulated in tumors compared to normal lung tissues, while *CCL4L2* was downregulated in tumors (all P < 10^-5^, permutation tests, **Figure 3F**). *DCAF8L2* showed limited expression and no significance in our analysis. Thus, *ATRNL1*, *CEP192*, and *VPS50* emerged as recurrence-associated candidate genes with mutational enrichment in R tumors and elevated tumor-specific expression, whereas *CCL4L2* was more frequently mutated in NR tumors and had lower expression in tumors compared to normal tissues.

### Pathway analysis implicates DNA repair processes and cilium organisation in lung tumor recurrence

Gene-focused mutational analyses may not fully capture complex biological mechanisms underlying tumor recurrence and less-frequently altered genes may remain undetected due to limited statistical power. To address these challenges, we mapped biological processes and molecular pathways that were preferentially mutated in R or NR tumors using the ActivePathways method [19]. Pathway enrichment analysis highlighted nine enriched pathways in R tumors and four pathways in NR tumors (family-wise error rate (FWER) < 0.1; **Figure 4A, Table S6, Figure S2**). Mutations in R tumors often occurred in DNA replication and double-strand break repair processes (e.g., GO:0031297 *replication fork processing*, FWER = 0.007), and cilium-related processes (e.g., GO:0044782 *cilium organisation*, FWER = 0.007). NR tumors were enriched in processes involving cell-cell adhesion and the nervous system (*e.g.,* GO:0007157 *heterophilic cell-cell adhesion*, FWER = 0.04). Pathway analysis also revealed an extended list of 54 candidate genes that had mutational enrichments in either R or NR tumors (P < 0.1, **Table S7**).

**Figure 4.**
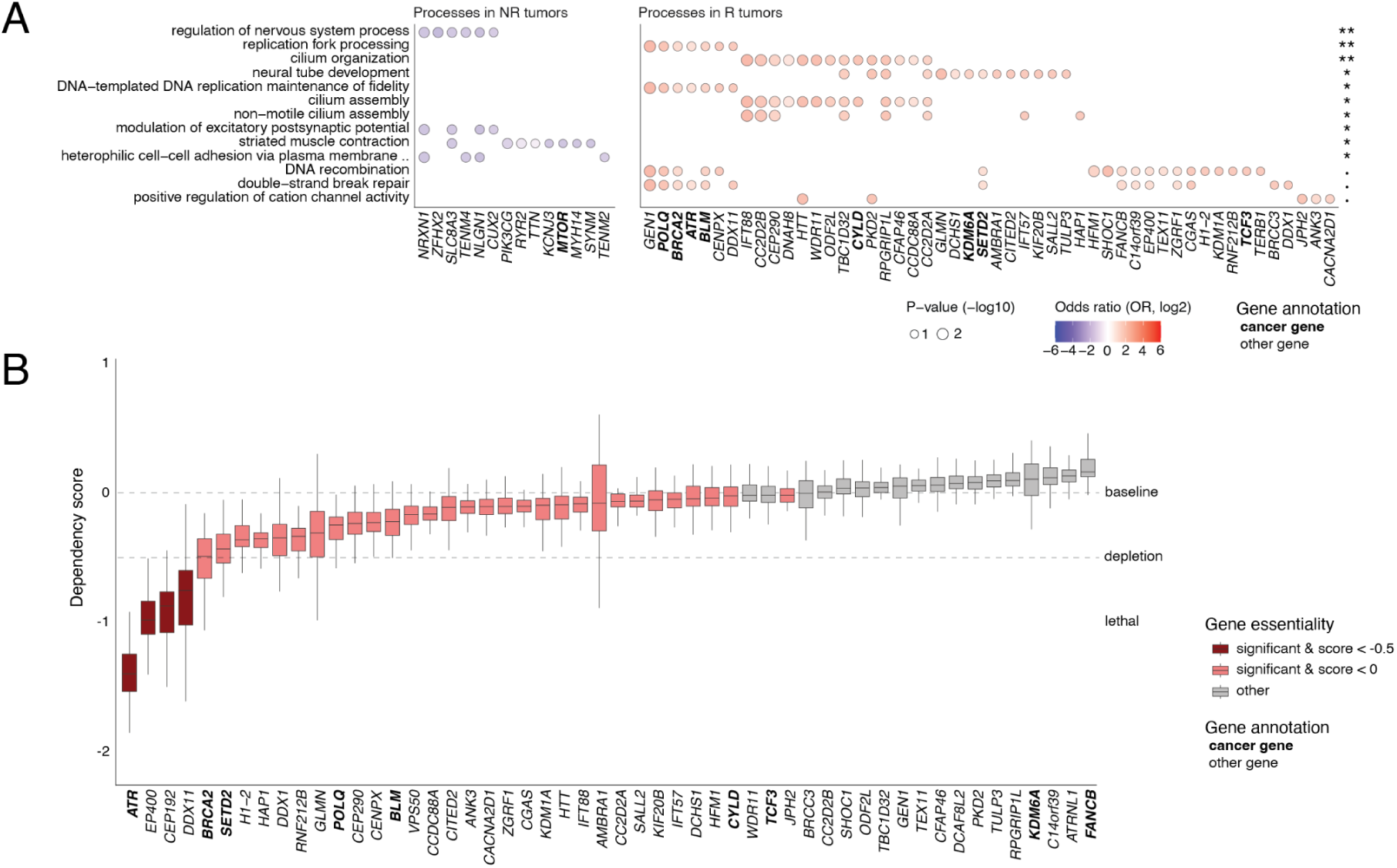
Biological pathways and essential phenotypes of candidate genes associated with lung cancer recurrence. **A.** Pathway enrichment analysis reveals biological processes and molecular pathways associated with tumor recurrence (R, recurrence; NR, no recurrence). The dot plot shows enriched pathways (Y axis) and corresponding pathway genes having mutational enrichments in R or NR tumors (X axis). Dot size corresponds to gene-based mutational enrichments. On the right, pathway-level adjusted P-values from ActivePathways are shown (Holm family-wise error rate, FWER). Asterisks indicate statistical significance ( • FWER < 0.1, * FWER < 0.05, ** FWER < 0.01, *** FWER < 0.001). **B.** Essentiality scores of candidate genes based on knockout screens in NSCLC cell lines in the DepMap project. Dependency scores (DS) of candidate genes were analysed for strong essentiality (DS < 0.5; dark red) or potential essentiality (DS < 0.0; pink). FDR-adjusted P-values from one-sample Wilcoxon tests are shown. Non-essential genes are shown in grey (FDR > 0.05). Established cancer genes are printed in boldface text.

Pathway analysis of recurrent tumors highlighted alterations in DNA replication and repair processes and the tumor suppressor gene *BRCA2*. When including hypermutated tumors, *BRCA2* mutations were nominally associated with tumor recurrence (11 R *vs.* 3 NR tumors, P = 0.02, one-tailed hypergeometric test). Three nonsense SNVs in *BRCA2* were found in hypermutated tumors that are either annotated as pathogenic or alter critical protein domains involved in DNA repair (p.W1692X, p.Y91X, p.Q3227X; **Table S3**). Another pathway gene *ATR* showed a trending association with tumor recurrence (8 R vs. 3 NR tumors, P = 0.065). Overall, these findings support DNA repair pathways and *BRCA2* as contributors to lung cancer recurrence.

Lastly, we evaluated the functional and therapeutic relevance of the candidate genes and pathways we identified. By analysing gene essentiality screens from NSCLC cell lines in the DepMap project [22], we found that the majority of our candidate genes (30/54) showed evidence of essentiality in NSCLC cells, including four found as strongly essential (*ATR*, *EP400*, *CEP192*, *DDX11*) (**Figure 4B**, **Table S8**). Examining established therapeutic associations, three genes *ATR*, *BRCA2*, and *KDM6A* are predictive of therapy response according to the OncoKB database [23]. Collectively, the genes and pathways uncovered in our analysis not only reveal potential mechanisms of recurrence but also point to clinically actionable targets to mitigate it.

## DISCUSSION

Here we studied primary lung tumors to identify candidate genes and pathways associated with disease recurrence. Elevated mutation burden and specific mutational processes appeared as the major characteristics of tumors with subsequent recurrence. High mutation burden has been linked to poor prognosis in NSCLC [27]. Also, the reactive oxygen species signature SBS18 that we associated with tumor recurrence is often found in lung tumors of never-smokers [28] and generates loss-of-function mutations in tumor suppressor genes [29]. We also prioritised certain genes and biological processes that may drive tumor recurrence or demarcate high-risk primary tumors, many of which have functional evidence in cancer cell lines or demonstrated therapeutic relevance, providing leads for future studies of lung cancer recurrence.

Our analysis suggests genomic hypermutation as a potential indicator of future tumor recurrence, a finding that has not been previously reported in lung cancer to our knowledge. Hypermutated tumors in our cohort were explained by mutations in DNA polymerase or mismatch repair genes, highlighting a link between defective DNA replication or repair and the hypermutated phenotype, in agreement with previous studies [24,25]. As high mutation burden is an established biomarker of immune checkpoint inhibition therapy [30], it is tempting to speculate that some patients at risk of lung tumor recurrence could benefit from immunotherapy.

To identify gene-level associations with tumor recurrence, we first examined established driver genes of NSCLC. While none of the known drivers were indicative of future recurrence in our cohort, we found that combined *TP53* and *CDKN2A* mutations were more common in NR tumors, potentially representing a lower-risk subset of lung cancers. *TP53* and *CDKN2A* encode key tumor suppressors in NSCLC [2,9,10] and their alterations have been associated with patient outcomes previously, including *TP53* alterations to improved prognosis and *CDKN2A* alterations to worse prognosis [31]. Thus, the recurrence associations of this potential genetic interaction require validation in larger cohorts.

Exome-wide analysis identified *ATRNL1* as the top recurrence-associated gene. *ATRNL1* is downregulated in cervical cancer and its overexpression was found to inhibit cell migration and epithelial-mesenchymal transition [32]. *ATRNL1* was implicated in lung cancer in one case report showing its insertion into the *EML4-ALK* fusion gene [33]. The locus also encodes the circular RNA gene *circATRNL1*, which is involved in metastasis and treatment response in ovarian and oral cancers [34,35]. Circular RNAs are increasingly recognised as post-transcriptional regulators of cancer initiation and progression [36]. Another recurrence-associated gene *CCL4L2* involved in inflammatory and immunoregulatory processes displayed a mutational hotspot causing recurrent G43R amino acid substitutions, an established mutational pattern of oncogenic activation [37]. Lastly, *CEP192* encodes a key centrosomal and cell cycle protein. Thus, our top candidate genes are potentially involved in hallmark cancer processes, providing leads for validation experiments.

Pathway enrichment analysis associated tumor recurrence with alterations in DNA replication and repair processes including *BRCA2*. *BRCA2* alterations in lung cancer are less prevalent, although studies have reported rare pathogenic germline variants [38]. Germline alterations of *BRCA2* sensitise tumors to PARP inhibition therapy through a synthetic lethality mechanism [39]. Tumor recurrence was also associated with mutations in cilium-related pathways. Primary cilia are microtubule-based organelles on cell surfaces that function in cell motility and signal transduction [40]. Cilia are lost in certain cancers, whereas other cancer types rely on cilia for growth and survival. Thus, further examination of DNA repair pathways and cilia in the context of lung tumor recurrence may generate mechanistic and therapeutic insights.

Limitations of our study include the lack of reference germline genomic profiles and cohort size. While most germline variants were filtered carefully using a panel of normals, our variant calls still likely contain a mix of tumor-specific somatic variants and rare germline variants with both benign and pathogenic effects. Sample size of our cohort limits the statistical power of our analyses and may reduce the generalizability of results. We only studied small mutations including SNVs and indels while copy-number alterations and structural variants were not considered. Therefore, additional genomic variants associated with tumor recurrence remain to be detected in future studies. Despite these constraints, our study provides insights into potential genetic mechanisms and predictors of tumor recurrence in primary lung cancer, warranting further validation in larger cohorts.

## Acknowledgements

This work was supported by Canadian Institutes of Health Research (CIHR) Project Grants to J.R. (PJT-162410, PJT-197925), an Investigator Award to J.R. from Ontario Institute for Cancer Research (OICR), and a New Investigator Award Terry Fox Research Institute (TFRI) to J.R. This research was also supported by a research grant from North Estonia Medical Centre (NEMC) to K.O. (TAKUP192). This research was also supported by an Estonian Research Council grant to A.V.M. (PSG608) and the research infrastructure “ELIXIR Estonia” funded by the Estonian Research Council to A.V.M. (TARISTU24-TK4). L.L. acknowledges mobility funding from the Kristjan Jaak National Scholarship Programme of Estonia. A.T.B. was partially supported by the Ontario Graduate Scholarship (OGS). M.B. and K.C.L.C. were partially supported by Medical Biophysics fellowships from University of Toronto. We would like to thank all patients and their families for donating tumor samples that enabled this research project. The results shown here are in whole or part based upon data generated by the TCGA Research Network: https://www.cancer.gov/tcga.

## Author contributions

L.L. led data analyses and figure generation. A.V. led clinical data curation and interpretation. A.T.B. led variant calling and quality assessment analyses. M.B. contributed to variant analyses. K.C.L.C. and A.T.B. led mutation signature analyses. L.L., A.V. and J.R wrote the manuscript with input from all coauthors. A.U., A.V.M., A.T.B., and K.O. contributed to study design, data interpretation, and manuscript writing. A.V. and K.O. led cohort development and DNA sequencing efforts. J.R. supervised data analyses. K.O. supervised cohort development. J.R. and K.O. conceptualised and co-supervised the study. All co-authors edited and approved the final manuscript.

## Data availability

Raw sequencing data and complete variant calls are not shared due to limited patient consent. Processed datasets with anonymised patient information are shared as supplementary tables.

