## Supplementary Materials for "Genomic profiling implicates candidate genes and mutagenic pathways driving lung cancer recurrence"

#### SUPPLEMENTARY METHODS

Study Population. Patients (n = 155) were selected from the North Estonia Medical Centre (NEMC) Thoracic Oncology Database. The main group of patients were selected based on diagnosis of NSCLC recurrence between 1 January 2015 and 31 December 2017 (referred to as the R group, n = 72). The control group included patients who did not develop recurrence (referred to as the NR group, n = 83). The NR group was selected to match with the R group by the type of previous treatment (surgery, no surgery), histology (squamous cell carcinoma, adenocarcinoma, others), TNM stage (version 7), age, sex, and follow-up time from the primary treatment. As follow-up since diagnosis, time to recurrence was reported in the R group (median 1.5 years, 95% range 0.13—7.6 years) and right-censored overall survival in the NR group (median 8.5 years, 95% range 4.4—17 years). All included patients were previously curatively treated. Disease recurrence was confirmed via imaging by computer tomography (CT). Clinical data was retrospectively collected from electronic health records. No patients received targeted or immunotherapy as these were not available for curative intent at the time. Histological specimens from formalin-fixed paraffin-embedded (FFPE) primary tumor samples originated from resection materials and biopsies at the time of NSCLC diagnosis. Only tumor tissue was analysed as no matched samples with germline reference were available. Samples were routinely processed and stained with hematoxylin and eosin to determine tumor subtypes. The study was approved by the ethics committee of the Estonian National Institute for Health Development on 20 September 2018 (approval # KK2453). Informed consent was waived by the ethics committee as the study was deemed to involve minimal or no risk to patients.

Whole-exome sequencing (WES). DNA was extracted from FFPE tumor samples using the Maxwell16 FFPE Plus Lev DNA Purification Kit in combination with the Maxwell® Lev instrument (Promega). DNA quantification was performed with the Quant-iT PicoGreen dsDNA Assay Kit (ThermoFisher). For each sample, 2 µg of DNA was fragmented using the LE220-plus system (Covaris). A 0.7x bead-based cleanup with AMPure XP size-selection beads (Beckman Coulter) was carried out to eliminate smaller DNA fragments. Fragmented DNA was quantified to ensure an input of 500 ng for library preparation. Libraries were generated using the KAPA HyperPrep Library Prep Kit (Roche) according to manufacturer's protocols. Library quality was assessed using the Fragment Analyzer DNA/NGS Kit (Agilent) and quantified with qPCR using the KAPA Library Quantification Kit (Roche). Libraries were pooled for exome capture with the xGen Exome Research Panel v1.0 (IDT), followed by qPCR quantification. Captured pools were normalised, combined based on qPCR concentration, and loaded for sequencing at 1.5 nM. Tumor samples were sequenced using 150-bp paired-end sequencing on the NovaSeq 6000 platform (Illumina) at Intermountain Precision Genomics (IPG), St. George, Utah, USA. The average sequencing coverage over the target region was 734X.

Genomic variant calling. We processed DNA sequencing reads for genomic variant detection and annotation, focusing on small mutations comprising single-nucleotide variants (SNVs) and small insertions-deletions (indels). WES data were processed using the Nextflow core Sarek pipeline (v3.4.2) executed with Nextflow (v24.04.3) under Singularity containerization [1,2]. FASTQ files for each tumor sample were processed in an independent Nextflow run without germline controls. Default parameters for Sarek read alignment and quality control were used. Briefly, raw reads were aligned to the human reference genome GRCh38 (GATK.GRCh38) using the BWA-MEM algorithm (v0.7.17-r1188). Aligned BAM files were processed using the GATK4 pipeline (v4.5.0.0) that was configured for duplicate marking (MarkDuplicates), base recalibration (BaseRecalibrator and ApplyBQSR), and contamination estimation (CalculateContamination) [3]. To maximise sensitivity and specificity for different variant classes, a multi-caller strategy was used. Variants were called using three independent software tools Mutect2 (GATK v4.5.0.0), Strelka2 (v2.9.10), and FreeBayes (v1.3.6) [4–6]. For increased confidence, we filtered variants having low support or low mapping quality according to variant calling pipelines. Specifically, we removed SNVs and indels that were not flagged as “PASS” in Mutect2 and Strelka2 VCFs and variants with a minimum “QUAL” score below 80 in FreeBayes VCFs. As germline variants were not available in this cohort, we used a panel-of-normals approach to filter the most likely germline variants from our dataset using the population genomics database gnomAD. Variants with major allele frequency (MAF) above 1% in ExAC03 non-TCGA, gnomAD exome (v4.0), and gnomAD genome (v4.0) were excluded from analysis [7,8]. Exome-wide sequencing of tumor samples identified 1,497,758 SNVs and 26,440 indels. The panel-of-normals filter excluded germline variants and revealed 184,913 SNVs and 3043 indels as likely somatic variants representing the tumor genome, corresponding to a 88% reduction of original variant calls. High-confidence variants were annotated with the ANNOVAR software (version 2020Jun08) [9] to obtain protein-coding impact and functional impact predictions of SNVs and indels.

Analysis of clinical covariates and mutation burden. Standard statistical tests were used to compare the context of clinical characteristics and tumor mutation burden of R and NR tumors ($P < 0.05$). To compare clinical stages between R and NR tumors, two-sided Wilcoxon rank-sum tests were used. Two-tailed hypergeometric tests were applied to compare histology types between R and NR groups. To compare mutation burden in R and NR groups, we counted variants in each tumor sample in three ways (SNVs, indels,

SNVs+indels). Variant counts were compared between R and NR tumors using two-sided Wilcoxon rank-sum tests. Next, we classified a subset of tumors as hypermutated based on considerably higher overall mutation burden (more than 3000 SNVs or indels per tumor). This custom approach was used because the standard hypermutation threshold (10 protein-coding mutations per Mbps) [10] is not applicable here since rare germline variants also contributed to overall mutation burden.

Associations with polymerase and mismatch repair deficiencies. To explore potential mechanisms underlying hypermutation in our cohort, we examined two major pathways involved in somatic hypermutation: DNA polymerase (POL) genes (*POLE*, *POLD1*, *POLQ, POLE2*) and mismatch repair (MMR) genes (*MSH2*, *MSH6*, *MLH1*, *PMS2*). First, tumor samples were annotated based on the presence of protein-coding mutations in POL or MMR genes. Second, distributions of POL or MMR mutations in R or NR tumors, and mutation burdens for SNVs and indels were compared. Differences in mutation burden were evaluated using two-sided Wilcoxon rank-sum tests and two-tailed hypergeometric tests were used to evaluate whether POL or MMR mutations were associated with R or NR groups. Mutations were annotated functionally and clinically using the ANNOVAR software and the ClinVar database [11].

Mutational signature analysis. We analysed mutational signatures of single-base substitutions (SBS) in R and NR groups. SBS signatures were decomposed and SNVs were ascribed to specific mutational processes using the SigProfiler family of methods [12]. SigProfiler was used to estimate the contribution of SBS signatures in each cancer sample using signature definitions from the COSMIC database (V3) [13]. The python package SigProfilerAssignment with the function Analyze.cosmic_fit was run on the filtered and merged consensus VCF files with the parameter “exome” set to ‘True’ and the “GRCh38” as the reference genome. Each tumor exome was run independently by fitting SBS signatures to the filtered VCF file. Only single base substitution (SBS) signatures with a total count greater than 1000 mutations across the cohort were analysed and artefact signatures were excluded (SBS27, SBS43, SBS45, SBS46, SBS47, SBS48, SBS49, SBS50, SBS51, SBS52, SBS53, SBS54, SBS55, SBS56, SBS57, SBS58, SBS59, SBS60, SBS95). For each tumor sample, the total number of mutations attributed to each SBS signature was calculated and signature contributions were normalised as percentages of the total per sample. Two-sided Wilcoxon rank-sum tests were used to compare signature contributions between R and NR tumors, followed by multiple testing correction using the Benjamini-Hochberg false discovery rate (FDR) method [14] and selection of significant results (FDR $< 0.05$).

Associating gene mutations with tumor recurrence. We analysed protein-coding mutations in individual genes in R and NR tumors to determine associations with tumor recurrence. After excluding hypermutated samples as defined above, a total of 149 tumor samples were included in subsequent analyses (67 R samples and 82 NR samples). Only protein-coding SNVs and indels based on Annovar annotations were used (non-synonymous, stop-gain, start-loss, stop-loss, frameshift, non-frameshift). Statistical analysis for gene-focused mutational enrichments was performed using two-tailed hypergeometric tests, P-values were adjusted using FDR and significant genes were selected (FDR < 0.05). First, we first assessed mutations in a well-defined set of driver genes in NSCLC identified in the TCGA PanCanAtlas project [15]. We selected 31 of 35 genes that were found in either LUSC or LUAD and had at least one protein-coding mutation in our dataset. Next, we tested combined mutations in the two genes that showed sub-significant trends in the previous analysis (*TP53* and *CDKN2A*) and revealed an enrichment in the NR group. Finally, we expanded the analysis to all protein-coding genes, excluding genes with mutations present in fewer than ten samples to reduce multiple testing penalties. Using hypergeometric tests, we classified genes as significant (FDR < 0.05), nominally significant (P < 0.01; i.e., second-tier), and non-significant (P > 0.01). To evaluate mutations in the context of protein sequence, we retrieved protein domain information from the InterPro database [16].

<u>Pathway analyses.</u> To identify biological processes and molecular pathways in differentially mutated genes in R or NR tumors, we performed pathway enrichment analysis using the ActivePathways method and standard protocols [17,18]. Pathway analysis was performed on gene P-values from the mutational enrichment analysis described above, and separate analyses were conducted for genes more frequently mutated in R or NR tumors. Biological processes from Gene Ontology [19] and pathways from the Reactome database [20] were downloaded from the g:Profiler web server [21] on April 16th 2024. Only gene sets containing between 25 and 500 genes were considered. The background gene list included all protein-coding genes. Multiple-testing correction and filtering was conducted using the default approach in ActivePathways (Holm family-wise error rate (FWER)) and significant pathways were selected (FWER < 0.1).

<u>Functional evaluation of candidate genes.</u> Candidate genes from gene-focused and pathway analyses were further evaluated in the context of gene expression in lung tumors and healthy tissues, gene essentiality screens in NSCLC cell lines, and known therapeutic targets. To study candidate gene expression in normal lung tissues and lung tumors, processed transcriptomics (RNA-seq) datasets from TCGA PanCanAtlas [22] and normal lung tissues in Genotype-Tissue Expression (GTEx) [23] projects were used. Transcriptomes of normal lung tissues were obtained from the GTEx Data Portal (gene_tpm_v10_lung.gct.gz). NSCLC transcriptomes were obtained for LUAD and LUSC (EBPlusPlusAdjustPANCAN_IlluminaHiSeq_RNASeqV2.geneExp.tsv) from Genomics Data Commons. To jointly analyse these two large and distinctly processed transcriptomics datasets, we first filtered the datasets to a common set of protein-coding genes. Next, we jointly normalised the two datasets by ranking each gene in each

sample from the lowest to the highest. To evaluate differential expression in lung tumors relative to normal lung tissues, we derived mean expression ranks for each gene in both tissue types and computed the fold-change (FC) values (tumor *vs.* normal). Custom permutation tests were used to evaluate significance of FC values, in which tumor and normal tissue ranks were randomly shuffled to derive random gene FC values over 100,000 iterations. Empirical P-values comparing true FC values relative to randomly-derived FC values were computed and significant results were selected ($P < 0.05$). To obtain functional evidence of prioritised genes in NSCLC cell lines, essentiality scores of candidate genes from gene-level and pathway-level analyses were retrieved from the DepMap project [24] comprising gene enrichment scores from CRISPR-based screens (DepMap Public 24Q4+Score, Chronos). For each gene, we asked whether dependency scores were significantly less than 0 (potentially essential) or less than -0.5 (strongly essential) using one-tailed one-sample Wilcoxon signed-rank tests with corresponding reference values, followed by FDR correction. Lastly, we queried the OncoKB database [25] to explore therapeutic associations of candidate genes.

### SUPPLEMENTARY FIGURES

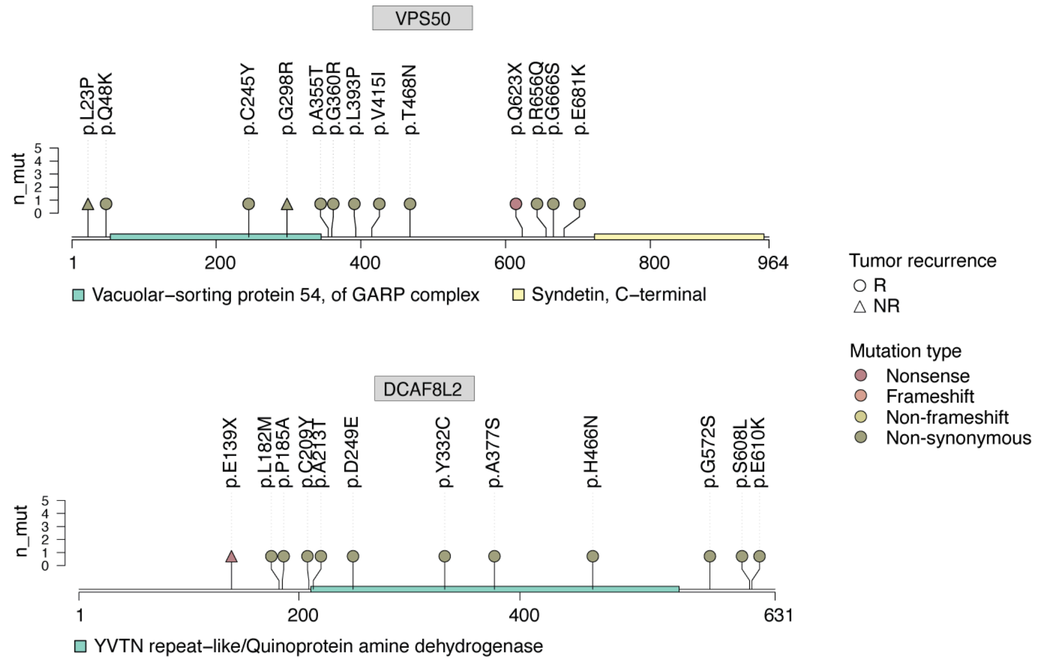


**Figure S1. Mutations in candidate genes *VPS50* and *DCAF8L2*.** Lollipop plots indicate the locations and affected protein domains of mutations in *VPS50* and *DCAF8L2*. Shapes on the Y axis indicate recurrence outcomes of mutated tumors (R, circle; NR, triangle) that are colored by protein-coding impact of these mutations.

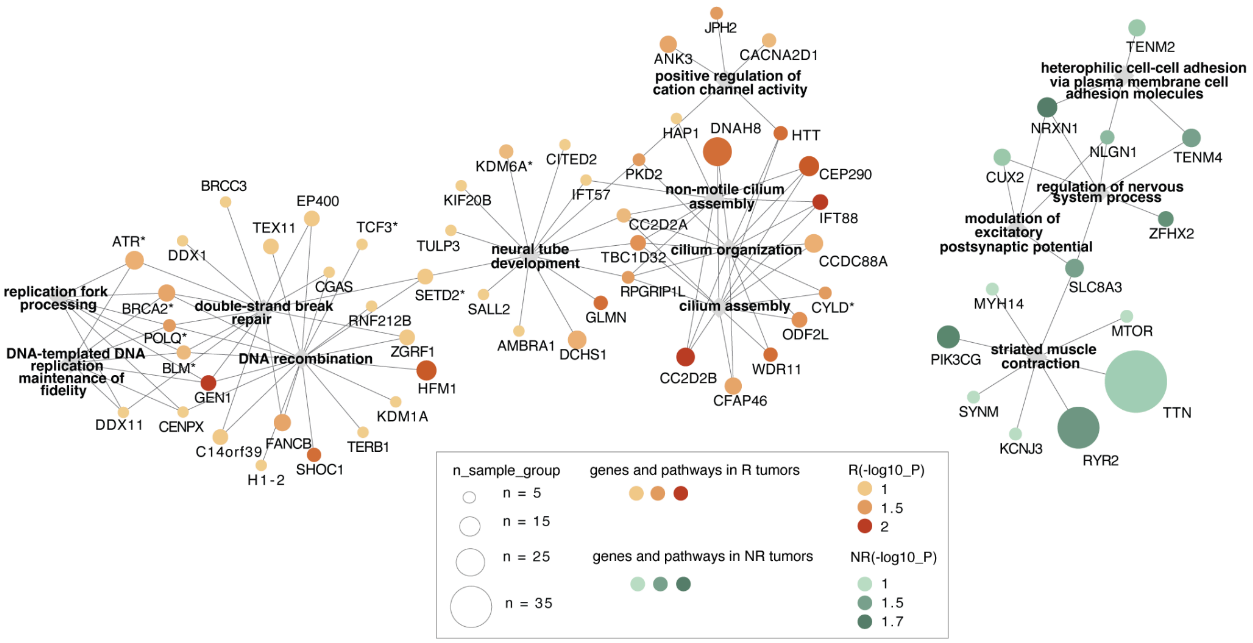


**Figure S2. Network view of pathways and genes enriched in mutations in R and NR tumors.** Interaction network of enriched pathways (shown as diamonds) reveals additional frequently mutated genes involved in each pathway (shown as circles) that were associated with recurrent (R) or non-recurrent (NR) outcomes in NSCLC. The size of each circle represents the number of tumor samples with a given gene mutation in either R or NR group. Established cancer genes are marked with an asterisk.
